# Longitudinal caries prevalence in a comprehensive, multi-component, school-based prevention program

**DOI:** 10.1101/2020.10.22.20217760

**Authors:** Jacqueline R. Starr, Ryan R. Ruff, Joseph Palmisano, J. Max Goodson, Omair M. Bukhari, Richard Niederman

## Abstract

**Background:** Globally, children’s caries prevalence exceeds 30% and has not markedly changed in 30 years. School-based caries prevention programs may be an effective method to reduce caries prevalence, obviate traditional barriers to care, and use aerosol-free interventions. The objective of this study was to explore the clinical effectiveness of a comprehensive school-based, aerosol-free, caries prevention program.

**Methods:** We conducted a 6-year prospective open cohort study in 33 U.S. public elementary schools, providing care to 6,927 children in communities with and without water fluoridation. Following a dental examination, dental hygienists provided twice-yearly prophylaxis, glass ionomer sealants, glass ionomer interim therapeutic restorations, fluoride varnish, toothbrushes, fluoride toothpaste, oral hygiene instruction, and referral to community dentists as needed. We used generalized estimating equations to estimate the change in the prevalence of untreated caries over time.

**Results:** The prevalence of untreated caries decreased by greater than 50%: from 39% to 18% in phase 1, and from 28% to 10% in phase 2. The per-visit adjusted odds ratio of untreated decay was 0.79 (95% CI: 0.73, 0.85).

**Conclusions and Practical Implications:** We show that a school-based comprehensive caries prevention program was associated with substantial reductions in children’s caries, supporting the concept of expanding traditional practices to include office- and community-based aerosol-free care.

## Introduction

Globally, in low-, medium-, and high-income countries, children’s caries experience exceeds 30% and has not markedly changed in 30 years.^1^ Between 1990 and 2010, the United States attepted to address the caries epidemic by increasing children’s Medicaid spending by over 300% (from $4.0 billion/year to $12.5 billion/year, adjusted for inflation)^2^ and the number of dentists by 22% (from 163,000 to 199,000).^3^ However, at the national level, these investments had little impact on children’s caries experience in primary (+0.6%, from 51.5% to 52.1%) or permanent teeth (−3.8%, from 21.2% to 17.4%).^4^

School-based caries prevention is one platform that may address the global and national caries burden. More than a dozen federal agencies and national organizations now recommended school-based caries prevention.^5^ Consequently, over the last 15 year period, there was a dramatic increase in the number of available school-based caries prevention programs.^6, 7^ However, when compared with the standard of care practiced in traditional dental offices, most school-based programs offer limited care. As well, school-based programs exhibit considerable variation in care type (e.g., screening only, or screening plus one or two preventive measures), care frequency (e.g., once or twice per year), care focus (e.g., specific grades or children’s age), or care for specific teeth (e.g., occlusal surfaces of only permanent first molars).^6, 7, 8-10^ More broadly, state practice acts and financial incentives support the overuse of office-based treatment and the underuse of school-based prevention.^8,12^ Furthermore, although there is considerable information on the efficacy of an individual preventive intervention in clinical trials, there is little to no information on implementation or clinical effectiveness of combined interventions in school-based practices.^9^

From 2004-2010, we conducted a comprehensive school-based caries prevention program that provided biannual treatments to prevent and arrest caries on all primary and permanent teeth in children. All care was provided by dental hygienists. Previously, we reported on the cost effectiveness of the program ^8-12^. The objectives of this study were to demonstrate that a comprehensive school-based care can obviate barriers to treatment and be clinically effective.

## Methods

The Forsyth Institute, Boston, MA Investigational Review Board approved this study. Reporting follows STROBE guidelines (see Supplemental Information 6).^13^ We previously reported the study rationale, calibration, selection of protocols, interventions, and six-month preliminary outcomes.^**14**^

### Schools, participants, clinical program, and data collection

We solicited school systems in suburban (Lynn), rural (Cape Cod), and urban (Boston) Massachussettes for participation. Six schools were initially enrolled in the cohort (Phase I), followed by an additional 27 schools (Phase II). We followed participants from Phase I for up to five years, and those from Phase II for up to three years.

All children attending a participating school were eligible to participate. The only exclusion criterion was the absence of informed consent and assent. The clinical team distributed and collected paper informed consent on electronically readable forms (Teleform, Cardiff Software, Vista, CA) to each participating school for guardian signatures. The informed consent forms requested the child’s sex, race, and ethnicity. Paper informed consents were then securely transmitted to a central repository and converted into an electronic dental record. We recorded and stored all clinical data on a proprietary electronic dental record software system (New England Survey Systems, Brookline, MA).

For each primary and permanent tooth, clinicians determined whether the tooth surface was decayed, missing, filled, sound, or sealed. Precavitated lesions were not scored as caries. Clinicians also recorded treatment by tooth surface (e.g., sealant, interim therapeutic restoration), or mouth (e.g., fluoride varnish). At the completion of data collection, de-identified data were securely uploaded to a Data Coordinating Center for data cleaning and verification.

### Interventions

Enrolled children received twice-yearly examinations and comprehensive caries prevention performed in the school by calibrated dental hygienists. Prevention included the provision of: prophylaxis, glass ionomer pit and fissure sealants (Fuji IX, GC America), glass ionomer interim therapeutic restorations on asymptomatic carious lesions (Fuji IX, GC America), fluoride varnish on all teeth (Duraphat or Prevident, Colgate-Palmolive), toothbrushes, fluoride toothpaste (Colgate Big Red, Colgate-Palmolive), and chairside brushing instruction. All children were referred to their own dentist, a local dentist, or to a community health center as needed for acute dental care (see Supplementary Material 1 for details). Teachers and school nurses were instructed to alert the clinical team if a child had any post-treatment problems. We did not monitor or request self-reported home toothbrushing.

### Statistical analysis

We longitudinally tracked subjects by matching on full name and date of birth. We numbered the visits for each child sequentially, regardless of time elapsed between visits, and also calculated the time elapsed between successive visits. In this analysis, we included only subjects whose age was at least five years at the initial visit, and we excluded any visits for subjects older than 12 years.

We excluded schools with fewer than 75 participating students (N= 16), subjects whose date of birth or age at study entry was missing (n=89 and n=151, respectively), and visits greater than 6 for a given subject (n=99). For the temporal trend analysis, we also excluded subjects with only one visit (n=1,625).

We divided the schools’ data analysis into two phases: Phase I included the first six schools; and Phase II included the remaining 27 schools with later program initiation. Phase I data included 2,588 subjects with 7,596 visit records. Phase II data included 4,339 subjects with 10,762 visit records.

From the dental examination and treatment, we derived indicators at the tooth- and child-level (e.g., untreated decay on any surface of any tooth) and the number of teeth with any untreated decay. We created these indicators separately for all primary and permanent teeth.

In the analytic set, we identified 333 visit records (out of 13,635) with at least one tooth identified as both permanent and primary. In these instances, the given tooth was included in analyses of permanent and primary teeth but counted only once in analyses that included both types of dentition. We also created indicators restricted to the occlusal surface of first molars.

At a child’s initial visit, we based the assessment of previous dental treatment on clinical examination. We also derived indicators for each child’s oral health status at their initial (baseline) visit: any untreated decay, caries experience (treated or untreated), and the number of teeth with untreated decay.

### Analysis of temporal trends in the prevalence of untreated decay

We used generalized estimating equations with a logit link and an exchangeable correlation matrix to evaluate the odds of untreated caries relative to the number of dental visits, up to 5 post-baseline visits in both Phases I and II. We omitted the baseline visit from analyses except by including an indicator of any untreated decay observed at baseline. In regression models, we included potential confounders, identified a priori as covariates associated with both the number of visits and dental outcomes and not influenced by either. These included: sex, previous dental treatment (yes/no), and age at examination (exact, in years, based on the date of birth and exam date; where exact dates were missing the day was assumed to be the 15^th^ day of the month). While we did not include a covariate for community water fluoridation, sensitivity analyses explored confounding at the school level which would by definition include water fluoridation. We included school indicators in Phase I analyses as planned a priori. The large number of phase II schools prohibited the inclusion of indicators in Phase II analyses. Children’s race was missing for a large proportion of subjects. Sensitivity analyses indicated that neither school (Supplementary Material 2) nor race (Supplementary Material 3) were likely to have confounded the results. We performed all analyses with dentition defined four ways: all teeth, permanent teeth only, primary teeth only, and occlusal surface of first molars only.

Temporal trends were assumed to reflect the effectiveness of school-based prevention. The validity of this assumption depends on whether children who stay in the program longer are similar to those who have only one or a few visits. To address potential attrition bias, we reanalyzed data restricting to subjects with an equal number of visits (3, 4, 5, or 6, in separate analyses, Supplementary Material 4). We performed an additional series of sensitivity analyses to probe the robustness of results under different assumptions, examining whether results changed in four different subsets of data: restricting to subjects who had <4 teeth with any untreated decay at baseline, had <6 teeth with any treated or untreated decay at baseline, or were younger than eight years at baseline, or restricting to visit numbers fewer than six (Supplementary Material 4).

## Results

### Participant Demographics

Demographics were similar for Phase I and Phase II. Approximately half the subjects were girls, close to 60% were ≤7 years old, and approximately 5% were 11 years old at their first exam (Table 1). Among the 31% reporting race in both phases combined, 19% were black, and approximately half were white, with most of the remainder either Asian or reporting more than one race (Table 1). The average participation rate, per school, was approximately 15% and ranged from 10% to 30%.

**Table 1.** Baseline demographic characteristics among children receiving school-based dental oral health care.

|  | **Total**<br>**(33 schools)**<br>**N (%)** |  | **Phase I**<br>**(6 schools)**<br>**N (%)** |  | **Phase II**<br>**(27 schools)**<br>**N (%)** |  |
| --- | --- | --- | --- | --- | --- | --- |
| **Total** | 6927 | (100) | 2588 | (100) | 4339 | (100) |
| **Sex** |  |  |  |  |  |  |
| Female | 3496 | (50) | 1271 | (49) | 2225 | (51) |
| Male | 3338 | (48) | 1286 | (50) | 2052 | (47) |
| **Age** |  |  |  |  |  |  |
| 5 | 996 | (14) | 254 | (10) | 742 | (17) |
| 6 | 1579 | (23) | 669 | (26) | 910 | (21) |
| 7 | 1368 | (20) | 624 | (24) | 744 | (17) |
| 8 | 1169 | (17) | 501 | (19) | 668 | (15) |
| 9 | 909 | (13) | 335 | (13) | 574 | (13) |
| 10 | 555 | (8) | 130 | (5) | 425 | (10) |
| 11 | 283 | (4) | 62 | (2) | 221 | (5) |
| 12 | 68 | (1) | 13 | (1) | 55 | (1) |
| **Race** |  |  |  |  |  |  |
| Black or African-American | 393 | (6) | 172 | (7) | 221 | (5) |
| White | 1209 | (17) | 173 | (7) | 1036 | (24) |
| American Indian, Alaska<br>Native, Native Hawaiian,<br>Pacific Islander | 50 | (1) | 9 | (0) | 41 | (1) |
| Asian | 230 | (3) | 56 | (2) | 174 | (4) |
| More than one race | 278 | (4) | 81 | (3) | 197 | (5) |
| Missing | 4767 | (69) | 2097 | (81) | 2670 | (62) |

**Table 2.**
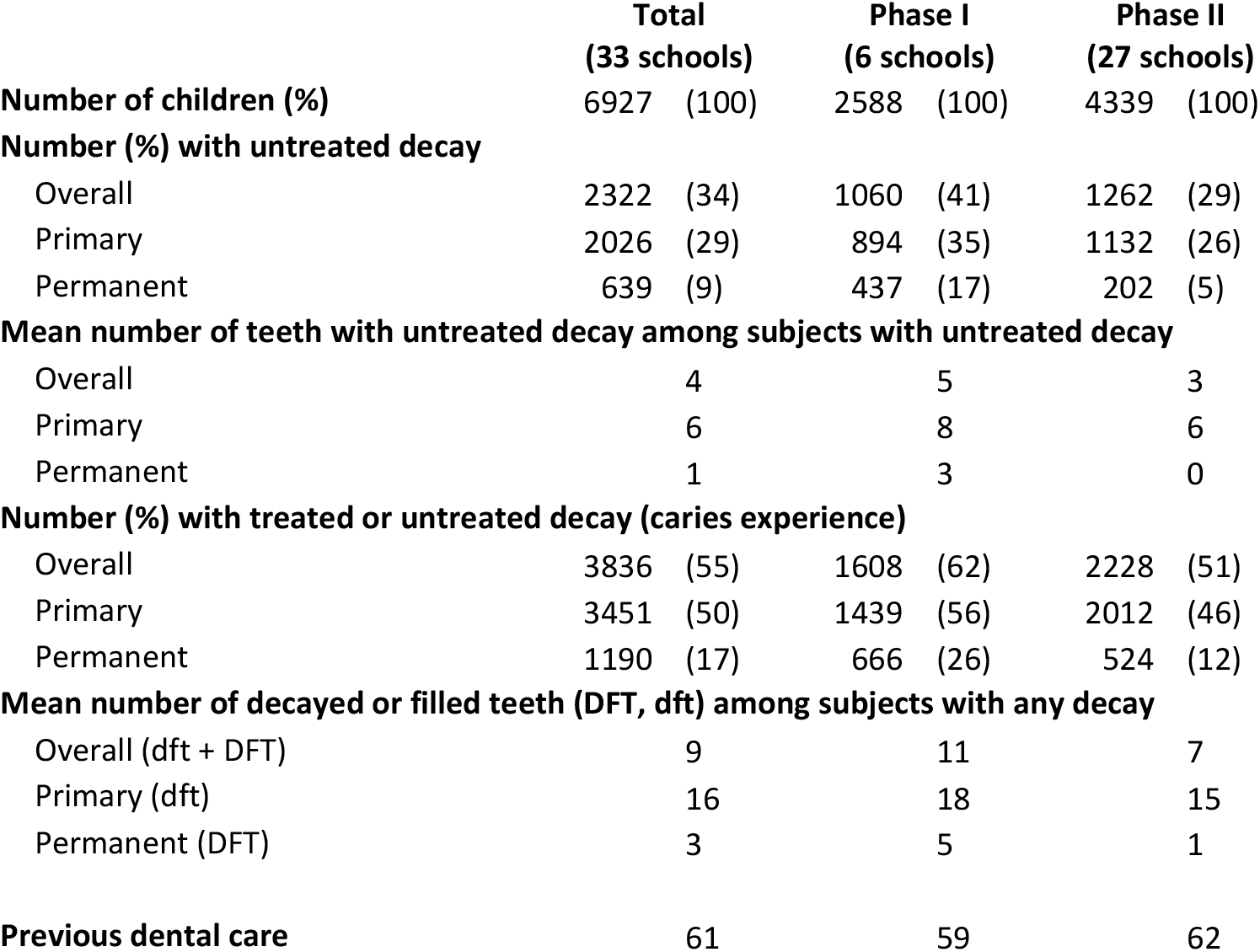
Baseline dental and oral health characteristics among children, aged 5 to 12 years, receiving school-based dental oral health care.

|  | **Total**<br>**(33 schools)** |  | **Phase I**<br>**(6 schools)** |  | **Phase II**<br>**(27 schools)** |  |
| --- | --- | --- | --- | --- | --- | --- |
| **Number of children (%)** | 6927 | (100) | 2588 | (100) | 4339 | (100) |
| **Number (%) with untreated decay** |  |  |  |  |  |  |
| Overall | 2322 | (34) | 1060 | (41) | 1262 | (29) |
| Primary | 2026 | (29) | 894 | (35) | 1132 | (26) |
| Permanent | 639 | (9) | 437 | (17) | 202 | (5) |
| **Mean number of teeth with untreated decay among subjects with untreated decay** |  |  |  |  |  |  |
| Overall | 4 |  | 5 |  | 3 |  |
| Primary | 6 |  | 8 |  | 6 |  |
| Permanent | 1 |  | 3 |  | 0 |  |
| **Number (%) with treated or untreated decay (caries experience)** |  |  |  |  |  |  |
| Overall | 3836 | (55) | 1608 | (62) | 2228 | (51) |
| Primary | 3451 | (50) | 1439 | (56) | 2012 | (46) |
| Permanent | 1190 | (17) | 666 | (26) | 524 | (12) |
| **Mean number of decayed or filled teeth (DFT, dft) among subjects with any decay** |  |  |  |  |  |  |
| Overall (dft + DFT) | 9 |  | 11 |  | 7 |  |
| Primary (dft) | 16 |  | 18 |  | 15 |  |
| Permanent (DFT) | 3 |  | 5 |  | 1 |  |
| **Previous dental care** | 61 |  | 59 |  | 62 |  |

**Table 3.** Estimated average per-visit change in odds of untreated decay among children enrolled in Title I schools in Massachusetts, receiving comprehensive oral health preventive care through the school-based program, by type of dentition.

|  |  | Phase I (six schools) |  |  | Phase II (33 schools) |  |  |  |
| --- | --- | --- | --- | --- | --- | --- | --- | --- |
|  |  |  | 95% CI<sup>a</sup> |  |  | 95% CI<sup>a</sup> |  |  |
| **Any baseline decay** | **n** | **OR<sup>b</sup>** | **upper** | **lower** | **n** | **OR<sup>b</sup>** | **upper** | **lower** |
| **All dentition** |  |  |  |  |  |  |  |  |
| No untreated decay at baseline | 1197 | 1.08 | 0.99 | 1.17 | 2356 | 1.04 | 0.97 | 1.12 |
| Any untreated decay at baseline | 776 | 0.79 | 0.73 | 0.85 | 936 | 0.74 | 0.68 | 0.80 |
| All subjects<sup>c</sup> | 1973 | 0.90 | 0.85 | 0.96 | 3292 | 0.88 | 0.83 | 0.93 |
| **Primary dentition** |  |  |  |  |  |  |  |  |
| No untreated decay at baseline | 1197 | 1.16 | 1.06 | 1.28 | 2356 | 1.10 | 1.01 | 1.20 |
| Any untreated decay at baseline | 776 | 0.90 | 0.83 | 0.97 | 936 | 0.76 | 0.70 | 0.82 |
| All subjects<sup>c</sup> | 1973 | 0.99 | 0.92 | 1.05 | 3292 | 0.90 | 0.85 | 0.96 |
| **Permanent dentition** |  |  |  |  |  |  |  |  |
| No untreated decay at baseline | 1197 | 0.80 | 0.69 | 0.94 | 2356 | 0.81 | 0.67 | 0.99 |
| Any untreated decay at baseline | 776 | 0.70 | 0.63 | 0.78 | 936 | 0.81 | 0.69 | 0.95 |
| All subjects<sup>c</sup> | 1973 | 0.73 | 0.66 | 0.80 | 3292 | 0.81 | 0.72 | 0.92 |
| **Occlusal surface first molar** |  |  |  |  |  |  |  |  |
| No untreated decay at baseline | 1197 | 0.85 | 0.70 | 1.03 | 2356 | 0.80 | 0.59 | 1.08 |
| Any untreated decay at baseline | 776 | 0.72 | 0.63 | 0.81 | 936 | 0.70 | 0.56 | 0.88 |
| All subjects<sup>c</sup> | 1973 | 0.74 | 0.67 | 0.83 | 3292 | 0.73 | 0.61 | 0.88 |

<sup>a</sup>CI=Confidence interval
<sup>b</sup>OR=odds ratio. Odds ratios were estimated by using a generalized estimating equations (GEE) approach with logit link, clustered on the subject, and are adjusted for sex, age at examination, and any previous dental care. Stratification by the presence of untreated decay at baseline was performed by including a multiplicative interaction term, visit times an indicator of any untreated decay at baseline.
<sup>c</sup>“All subjects” represents all subjects regardless of whether they had untreated decay diagnosed at the baseline visit, with results adjusted for the presence of untreated decay at baseline.

At their baseline visit, 33% of children had untreated decay in any dentition, with a range of 18% to 54% across the schools (Table 2). Dentition-specific decay prevalence at baseline was 29% for primary and 9% for permanent teeth. Fully 55% (across-school range: 28% to 68%) of children had caries experience (treated plus untreated decay). Subjects averaged 2.6 teeth with caries experience at baseline. These oral health indicators were slightly worse in Phase I subjects (Table 2). Approximately 20% had at least five teeth with caries experience at baseline, and 10% had at least three teeth with untreated decay at baseline (data not shown).

Of the 6,927 subjects in Phase I and II, 5322 (77%) had more than one visit. For Phase I, 45% of students had ≥3 visits; 33% had ≥4, and 18% had ≥5 visits. By definition, Phase II schools entered later in the study period and had a shorter duration in the program. For Phase II, 35% of students had ≥3 visits, 23% had ≥4, and 9% had ≥5 visits. In both study phases, the median time elapsed between visits was six months, and the 10^th^ and 90^th^ percentiles were approximately 4 and 13 months, respectively.

### Temporal Trend in Untreated Caries Prevalence

In Phase I, children with untreated caries decreased from 39% at baseline to 18% at visit 7 (54% reduction). In Phase II, children with untreated caries decreased from 28% at baseline to 10% at visit 7 (64% reduction). For children with untreated caries at baseline, the trends were similar in primary and permanent teeth and the occlusal surfaces of first molars for both Phase I and II (Figures 1 and 2).

**Figure 1.**
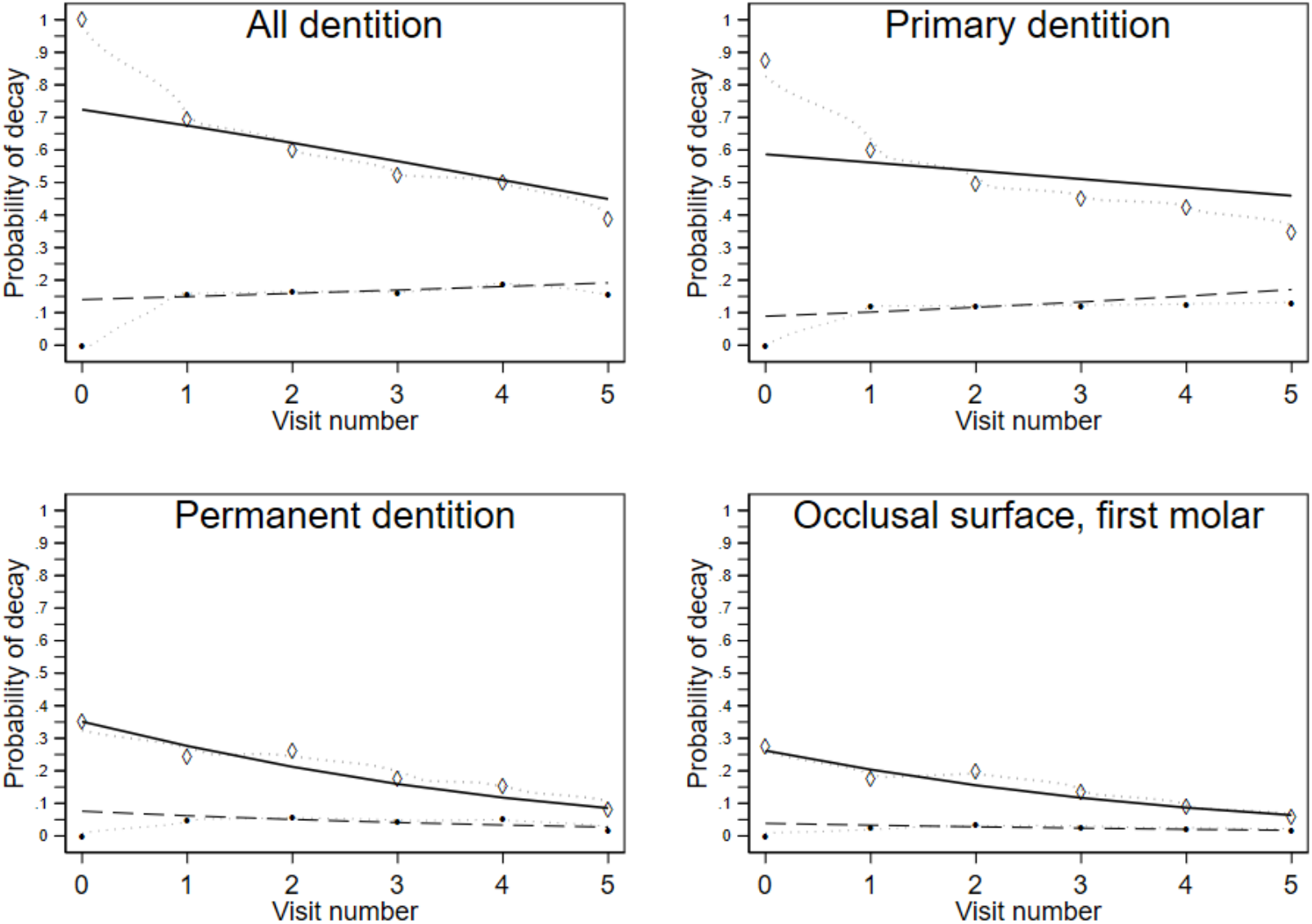
Observed (dashed curves) and predicted (solid lines) prevalence of untreated decay by visit number among children enrolled in study Phase I Title I schools in Massachusetts (n=6 schools), receiving comprehensive oral health preventive care, by type of dentition. Predicted prevalence was estimated from generalized estimating equations (GEE) approach with logit link, clustered on the subject, and are adjusted for sex, age at examination, and any previous dental care. Stratification by the presence of untreated decay at baseline was performed by including a multiplicative interaction term, visit times an indicator of any untreated decay at baseline. For subjects with untreated decay diagnosed at baseline, the dashed curve represents the observed prevalence by time point; the solid line represents the predicted prevalence. For subjects without evidence of untreated decay at baseline, the dashed curve represents the observed prevalence by time point; the solid line represents the predicted prevalence.

**Figure 2.**
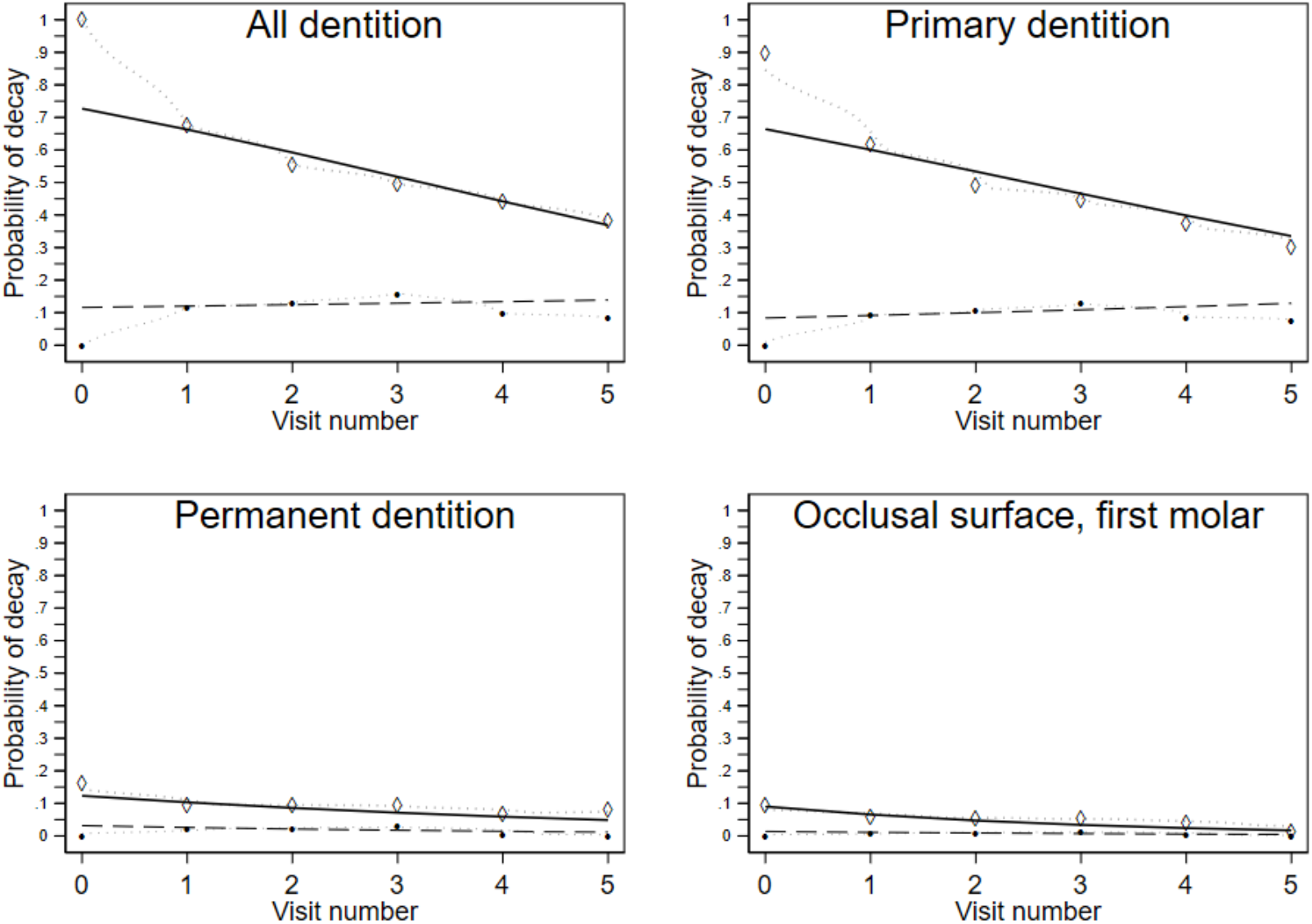
Observed (dashed curve) and predicted (solid line) prevalence of untreated decay by visit number among children enrolled in study Phase II Title I schools in Massachusetts (n=28 schools), receiving comprehensive oral health preventive care, by type of dentition. Predicted prevalence was estimated from generalized estimating equations (GEE) approach with logit link, clustered on the subject, and are adjusted for sex, age at examination, and any previous dental care. Stratification by the presence of untreated decay at baseline was performed by including a multiplicative interaction term, visit times an indicator of any untreated decay at baseline. For subjects with untreated decay diagnosed at baseline, the dashed curve represents the observed prevalence by time point; the solid line represents the predicted prevalence. For subjects without evidence of untreated decay at baseline, the dashed curve represents the observed prevalence by time point; the solid line represents the predicted prevalence.

Considering all children (with and without caries), and including both primary and permanent dentition, multivariable models indicated an average per-visit decrease in the odds of untreated caries (OR = 0.90, 95%, CI 0.85-0.96, Phase I; OR = 0.88, 95% CI 0.83-0.93, Phase II)(Table 3). The odds ratio was lower for the permanent dentition, and for the occlusal surface of permanent first molars and higher for the primary dentition (Table 3).

The per-visit change depended heavily on baseline dental health. Specifically, the most beneficial trend occurred in subjects who had any untreated decay at baseline (e.g., OR = 0.79; 95% CI 0.73, 0.85, Phase I) for primary and permanent dentition together (Figure 1 and Table 3). This trend was somewhat stronger for permanent teeth and specifically for the occlusal surfaces of first molars. In contrast, for children with no untreated decay at baseline, there was a slight upward trend, especially for the primary dentition (Figures 1 and 2). All point estimates were similar for Phase I and II analyses (Figures 1 and 2, Table 3).

### Other Analyses

In most of the sensitivity analyses, results were robust and offered the same interpretation as the primary analyses. There was no evidence that temporal trends in decay risk were due to confounding by the school (Supplement information 2, eTables 5a-b) or by race (Supplemental information 3, eTables 8a-c) or due to attrition bias (Supplemental information 4, eTables 11a-c). When we restricted subject analysis by age or based on the number of carious teeth at baseline, the results changed only minimally (Supplemental information 5, eTables 14a-c).

## Discussion

In this pragmatic study, we assessed the potential effectiveness of a multi-component, longitudinal, school-based caries prevention program delivered by dental hygienists. The program focused on U.S. schoolchildren attending Title 1^15^ elementary schools where > 50% of the student population participated in free or reduced lunch programs (a surrogate indicator for lower socioeconomic status).

The program was shown to reduce untreated caries by more than 50% over six visits. This 50% reduction is a change that, as explained below, would be very unlikely in the absence of school-based caries prevention. Furthermore, wide-ranging sensitivity analyses support the conclusions of a beneficial preventive effect. Parallel economic analysis of this cohort indicated that this program is both cost-saving and cost-effective when compared with no care or other prevention programs^12^, and the methods reported here offer one mechanism to expand the reach of traditional dental practices.^14^ These results also support the claim that a low-cost, high-access, community-based caries prevention program can control or reduce the prevalence of dental caries.^1,16^

The primary methodologic limitation of this study was the absence of a control group of children who did not receive care. Therefore, we tracked longitudinal trends in untreated caries as a surrogate for program effectiveness. This approach is valid if outcomes are similar for subjects who stay in the program throughout the follow-up period and for those who drop out of care. Reasons for shorter care duration include, for example, movement into or out of a school, administrative censoring, and late entry into the program (e.g., two visits in the last year of the program). We evaluated whether this attrition bias, rather than program effectiveness, explained the results. Specifically, we conducted a series of analyses in which we restricted the data to subgroups of subjects who all shared the same total number of visits. The conclusions regarding the effectiveness of caries prevention for permanent teeth were not altered, particularly for subjects who entered with existing untreated caries. Furthermore, if the program were ineffective, one would expect the odds of untreated caries to increase over time since tooth surfaces experienced more time at-risk for caries development. However, these increases did not occur.

Another limitation was the modest overall study participation based on the informed consent process.^17^ At the time we conducted this study, the informed consent process required a series of annual repetitive steps, including consent form delivery to the schools, teachers, students, and parents. Informed consent is, therefore, a significant challenge in all school prevention programs.^18, 19^ One concern is that parents who return consent forms may be among the most engaged, most economically secure, and most highly educated.^18, 19^ In the population studied here, the focus was Title 1 elementary schools with greater than 50% free or reduced lunch participation. In this context, it is notable that at baseline, 28% to 39% of children had untreated caries. These numbers are substantially higher than the current U.S. national average.^20^ Conversely, between 10% and 18% of the children ended the study with untreated caries. These averages are at or below current national averages.^4^ Extrapolating from the results in program participants, and assuming that children without signed consent forms likely had higher caries levels, they might have benefited even more than those with signed consent forms.

### Practical Implications

These results have several broad positive practical implications. First, and centrally, the findings demonstrate the feasibility and clinical benefit of one approach to school-based, caries prevention programs, delivered by dental hygienists. Second, the results support the contention that a nation-wide, comprehensive caries prevention program, implemented for all U.S. children, could reduce children’s caries by >50%, with potential cost savings of as much as half of what Medicaid currently spends for children’s oral health care.^8, 12^ Program support from Medicaid and insurers could expand care outside of the traditional dental practice.

### Clinical Implications

There are several broad clinical practice implications. First, we used a community risk assessment model to identify high-risk schools serving high-risk populations (>50% free or reduced lunch program).^21^ The fact that the baseline caries prevalence was well above national averages suggests that schools’ Title I designation was an appropriate criterion with which to identify groups of children at high risk of caries and with a high need for dental care. This approach contradicts current recommendations for individual caries risk assessment, radiographs, and teledentistry.^23-25^ Second, to improve health for all children, we selected and implemented multiple preventive interventions with evidence of effectiveness from systematic reviews.^9^ Our comprehensive approach contrasts with most school-based prevention programs that focus on specific teeth in specific age children and a limited number of preventive interventions.^6, 7^ The additional interventions add little cost to the program overall and can be performed quickly. Third, we used glass ionomer for sealing pits and fissures and for interim therapeutic restorations. The use of glass ionomers for these purposes differs from standard practice and guidelines recommending composite resin.^22^ Nevertheless, such use aligns with current systematic reviews.^23^ Fourth, the program practitioners did not remove caries before placing interim therapeutic restorations, which is harmonious with long-term clinical trials,^24, 25^ systematic reviews of efficacy,^26^ and American Dental Association clinical guidelines.

Given the preceding, it is notable that the program met all six of the Institute of Medicine’s quality criteria.^27^ Care was safe (∼1 in 2000 adverse events), effective (∼50% reduction in caries), patient-centered (care comes to children, rather than children coming to care), timely (care is delivered twice per year), efficient (all care takes less than 30 minutes), and equitable (all children with informed consent receive care, independent of their insurance or ability to pay). It is also notable that the program met health care’s triple aim of improving quality, improving health, and reducing costs.^28^ Finally, the program meets the U.S. Supreme Court’s standards of care definition ^29,30^, which are based on guidelines or systematic reviews of human randomized controlled trials published in peer-reviewed journals.

### Policy Implications

These results demonstrate the feasibility and clinical benefit of one approach to comprehensive, school-based caries prevention. This study included 7,037 students attending 33 multi-ethnic Title 1 Massachusetts elementary schools located in urban and rural areas, in areas with and without community water fluoridation, among children with and without caries, and among children from immigrant and non-immigrant families. Given the broad base of the participating population, the results should generalize to other populations.

### Aerosol-free preventive care reduces the risk of airborne disease transmission

The U.S. Occupational Safety and Health Administration (OSHA) categorizes dental care as very high risk ^31^ because of aerosols harboring and potentially transmitting bacteria, fungi, and viruses, such as SARS-CoV-2^32^ Virus laden aerosols in particular are detectable and viable for hours.^33^ Consequently, proximity to patients and the exposure to potential disease-borne aerosols place clinicians among workers with the highest infection risk.^34^ Based on OSHA definitions, infection risk can be reduced by removing the hazard (eliminating aerosol-based care) and replacing the hazard (providing aerosol-free care). Notably, the interventions provided as part of the program analyzed here are aerosol-free, and therefore reduce the OSHA defined risk by one category. These interventions and others, such as silver diamine fluoride, form a group of simple, aerosol-free, effective (SAFER) dentistry procedures. ^35^

### Barriers to Implementation

Despite this evidence, there remain several barriers to clinical and policy change. The economic and diffusion literature suggests that legislative and regulatory barriers for systematic implementation of caries prevention will be significant: studies indicate that 10-20% of stakeholders across governmental, organizational, clinical, and patient groups must support legislative, regulatory, or economic reform to effect wide-scale adoption of caries prevention. ^36, 37, 29^ Another barrier is that current Medicaid reimbursement rates cover neither the costs of care nor the value of care.^8,12^ Furthermore, most states require a prior examination or direct dental supervision before a hygienist provides care and do not allow hygienists to practice to the full extent of their training.^38^ Existing practice acts can, therefore, limit access to care due to the availability of dentists or their costs. When this program began, the Massachusetts state practice act required a prior dental examination. By study completion, the practice act allowed dental hygienists to assess needs and provide care with indirect supervision.

In sum, the results reported here support the concept that a comprehensive school-based caries prevention program can substantially reduce caries prevalence and meet the Institute of Medicines quality aims and health care’s triple aim. Widespread implementation could increase the reach of traditional dental practices while reducing the costs of care ^2, 39^ and inequity.^40^ Lastly, the use of aerosol-free procedures can provide safe dental care in pandemic environments, such as COVID-19.

## Data Availability

All data requests must be made in writing to the corresponding author.

## Acknowledgements

We thank the following individuals who contributed to the clinical and/or analytic work reported here: Alice Bisbee, Howard Cabral, MaryAnn Cugini, Ralph Kent, Ellen Gould, Denise Guerrero, Carolina Hommes, Timothy Martinez, Valarie Osborn, John Roberge, Aronita Rosenblatt, Karina Roldan, Jennifer Soncini, Michael Stanley, Philip Stashenko, Mary Tavares, Xiaoshan Wang, and Barbara Yates.

We thank the following community organizations who facilitated care for the children enrolled in the study: The Lynn and Harbor Community Health Centers and the Lynn, Boston, and Cape Cod school systems.

We particularly thank New England Survey Systems (J. Roberge, M. Stanley) for their work in developing the proprietary electronic patient record software used in data collection for this study. We also thank the Massachusetts Department of Health (M. Foley and L. Bethel) for background guidance.

This work was supported in part by the NIH/NIMHD: U24MDD006964

Finally, we posthumously thank and dedicate this paper to Dr. James Ware from the Harvard School of Public Health, who assisted us in developing the statistical analysis plan.

## Supplementary Material

**Supplemental information 1: Details about clinical examination**.

Dentists underwent visual/tactile technique calibration for dental caries (κ = 0.75) using the National Institute of Dental and Craniofacial Research diagnostic criteria as a standard reference.^1^ They dried tooth surfaces before examination without cleaning them or making radiographs.

Dental hygienists provided prophylaxis and oral hygiene instruction; provided toothbrushes and fluoride toothpaste; and placed glass ionomer sealants, glass ionomer temporary restorations (for carious teeth) and fluoride varnish, all based on the dental examination and treatment plan. The dentists and hygienists prepared written reports in parents’ and guardians’ native languages, through which they disseminated examination results and recommendations for treatment. Parents and guardians also received referrals to collaborating local dentists or community health centers if they did not have a dentist.

For emergency care, the clinical team followed the school protocol. The team first notified the school nurse, then the pupil’s parents. The collaborating dentists offered to set aside time to handle emergencies. Parents provided transportation. Nurses kept a log of post-treatment emergencies, had telephone numbers of local dentists and the program director to facilitate immediate care.

**Supplemental information 2: Assessment of confounding by the school**.

In the a priori data analysis plan, we expected that schools might confound the association between visit number and presence of untreated caries, in part because we had limited covariate information about individual subjects. As well, visit patterns and attrition rates might also vary for the subjects in different schools. For the six phase I schools, it was straightforward to perform explicit adjustment in regression models by including indicators for each school.

This same adjustment method was not feasible in phase II analyses. Phase II schools would have required 27 school indicators. We approached this limitation in two different ways. First, we sought empirical evidence of confounding, by the school, in the phase I analyses. We did this by fitting models with and without the school indicators and comparing the estimated association of untreated caries with visit number (and the standard errors for this association). The close similarity of the estimated coefficients and their standard errors (Table 2a) suggested there was little confounding by the school in the phase I analysis. Nevertheless, this does not guarantee that such a relationship did not exist in the phase II analyses, with 27 different schools.

Second, we performed both the phase I and phase II analyses by fitting multilevel mixed effects models with a random effect for school and fixed effects corresponding to other covariates. The results of these analyses were similar to the Phase I and II analyses using generalized estimating equations(GEE) (Table 2b). Compared with the GEE analyses unadjusted for school, the multilevel analyses with random effect for school yielded stronger odds ratio estimates for children entering the program with untreated dental caries.

**eTable 2a.**
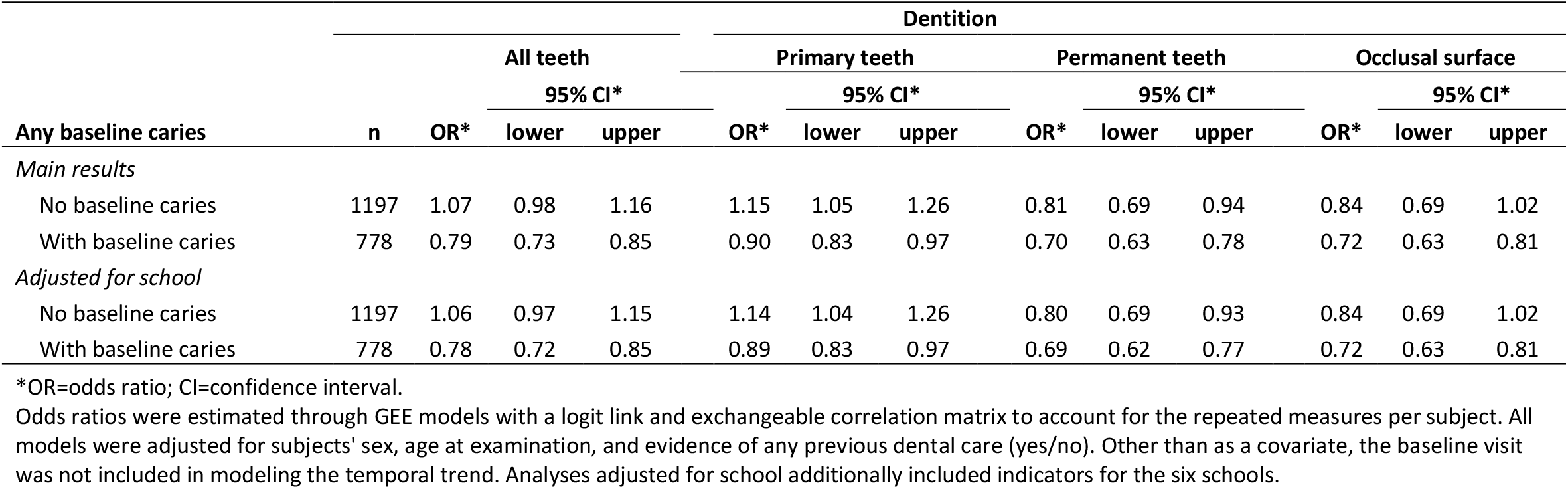
Temporal trend in the presence of untreated caries over visits: Additional adjustment for school in Phase I (first six schools).

**eTable 2b.**
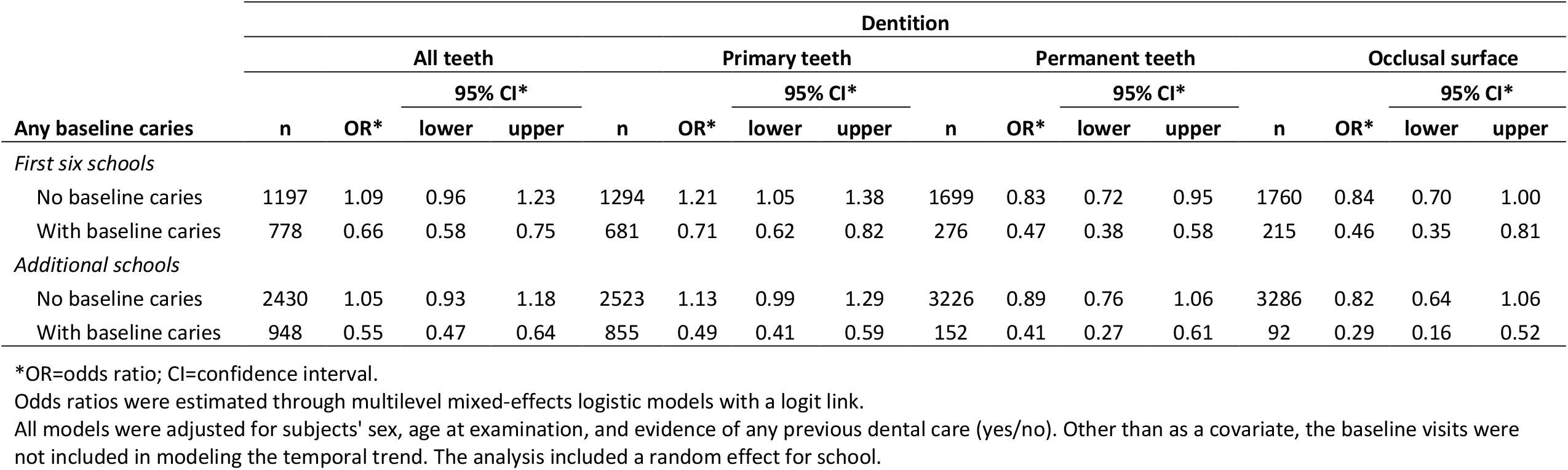
Per-visit change in prevalence of untreated caries among children: School random effects.

**Supplemental information 3: Assessment of confounding by race**.

Approximately 80% of the subjects were missing race. The non-white race is a risk factor for untreated caries and poorer oral health. There was not a specific reason to expect that visit patterns would also differ by race. Yet, to address this possibility, we first stratified the analyses according to whether subjects were missing race or not, without adjusting for race in either model. Second, we adjusted for race in the smaller proportion (∼20%) who reported race (Black; White; Asian, Native Alaskan, Native American, or Pacific Islander; or more than one race or unknown). We performed these analyses aggregating all the schools as well as separately for the first six (Phase I) and later 27 (Phase II) schools (Tables 3a, 3b, and 3c, respectively).

In most of these analyses, the odds ratio estimates of temporal trend differed only a small amount from the analyses of the main results. When the odds ratio estimates differed more substantially, it was generally to strengthen the results, not weaken them. These differences include apparent confounding by lack of reporting of race (or underlying characteristics associated with it).

Based on these sensitivity analyses, it appears that race was unlikely to have confounded the results in an anti-conservative direction.

**eTable 3a.**
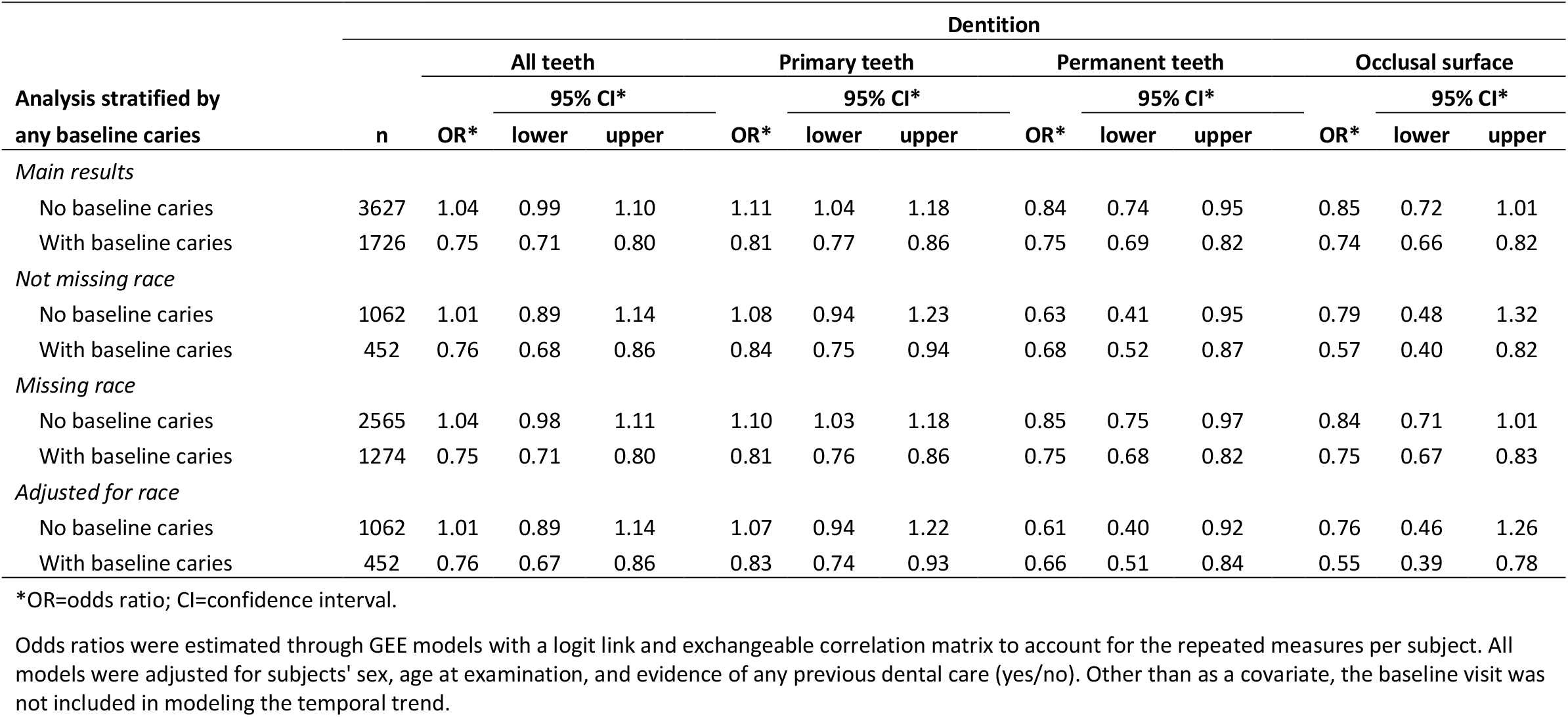
Temporal trend in the presence of untreated caries over visits: Exploration of possible confounding by race in all 33 schools (Phases I and II).

**eTable 3b.**
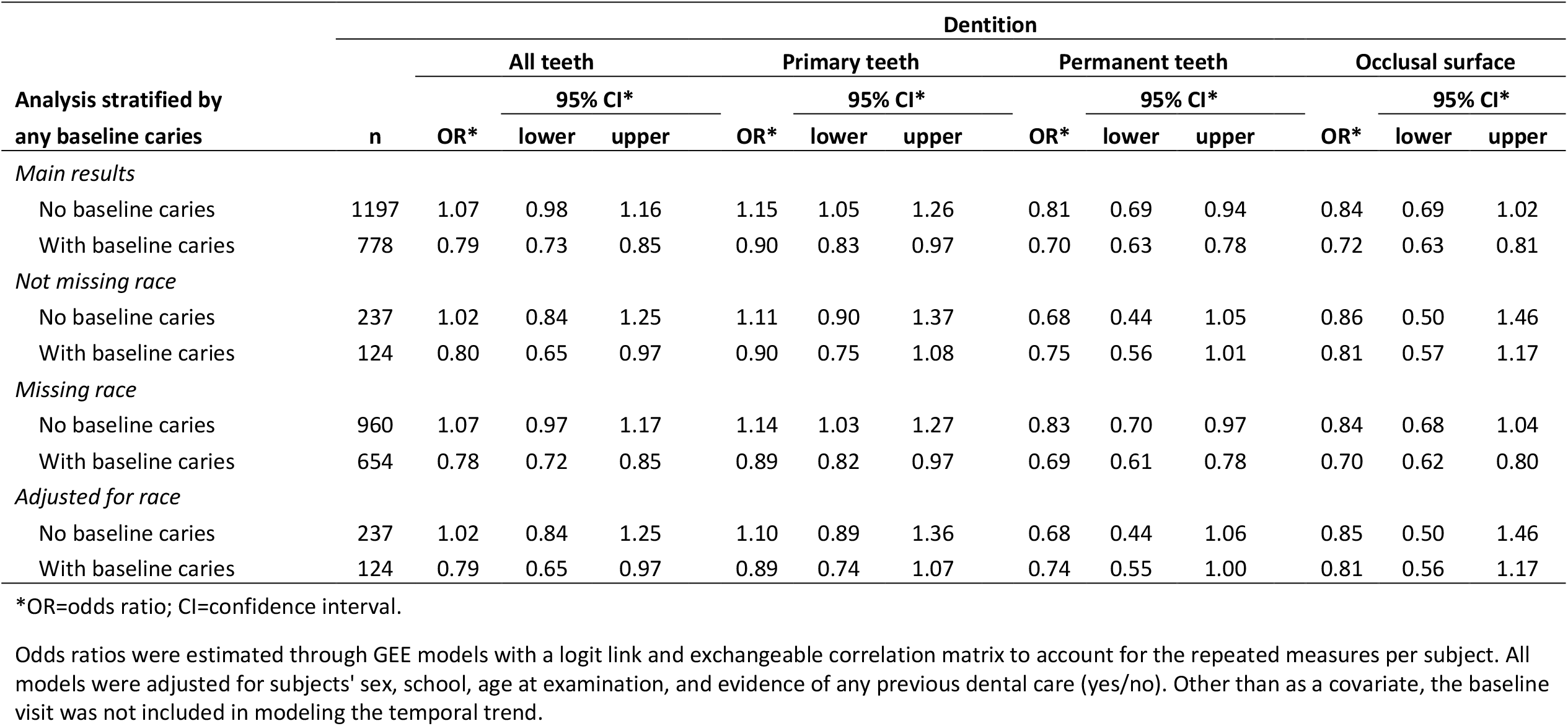
Temporal trend in the presence of untreated caries over visits: Exploration of possible confounding by race in first 6 schools (Phase I).

**eTable 3c.**
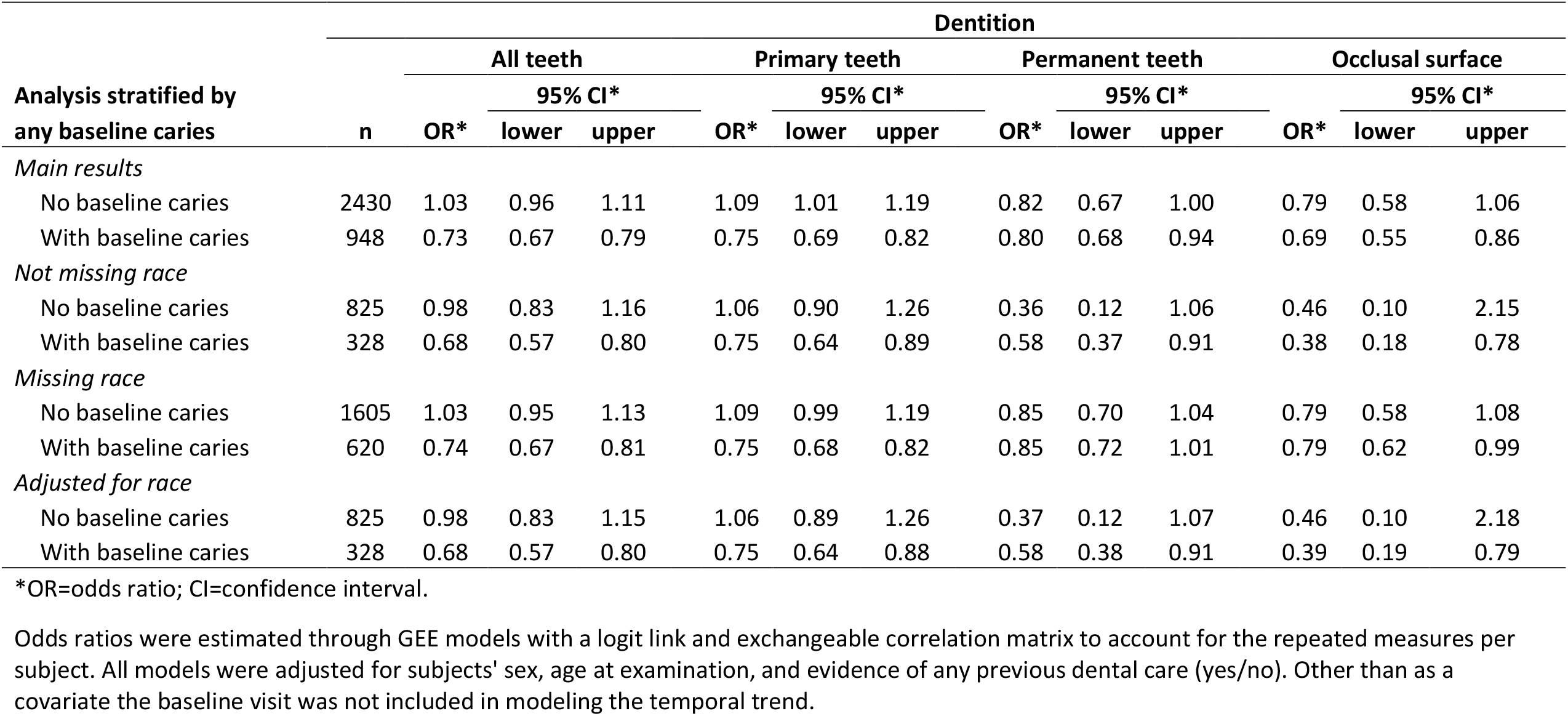
Temporal trend in the presence of untreated caries over visits: Exploration of possible confounding by race in latter 27 schools (Phase II).

**Supplemental information 4: Sensitivity analyses to address bias due to attrition**.

A significant limitation to this study was the lack of an untreated control group and the concomitant concern that subjects who had a longer duration of care, or a greater number of visits, differed in their underlying risk of untreated caries. If, for example, the subjects at highest risk of untreated caries preferentially dropped out after only two or three visits, the estimated decrease in odds of untreated caries over time could be overestimated.

To address this possibility, we fit a series of models in which subjects had a fixed total number of visits, either 3, 4, 5, or 6. We compared these results to the primary analyses for all schools (Table 4a), Phase I (6 schools, Table 4b), and Phase II schools (Table 4c).

In the subjects observed for 4, 5, or 6 visits, the estimated odds ratios were similar to those estimated from the whole population overall. Estimates of temporal trend changed most markedly among subjects who began with no caries at the baseline visit and had only three visits; among these subjects, the odds of caries increased over their three visits. The other group of subjects who had only three visits—those who began the program with caries diagnosed at baseline—experienced a similar decrease over time in the odds of untreated caries as the whole population overall, also similar to those observed for up to six visits.

We do not consider these differences to negate the study conclusions, because the population-averaged (adjusting for, but not stratifying by, the presence of caries at baseline) temporal trend is still negative even for subjects restricted to three visits. We know that over time, the risk of untreated caries increases. Furthermore, even effective caries prevention programs are not 100% effective.

**eTable 4a.**
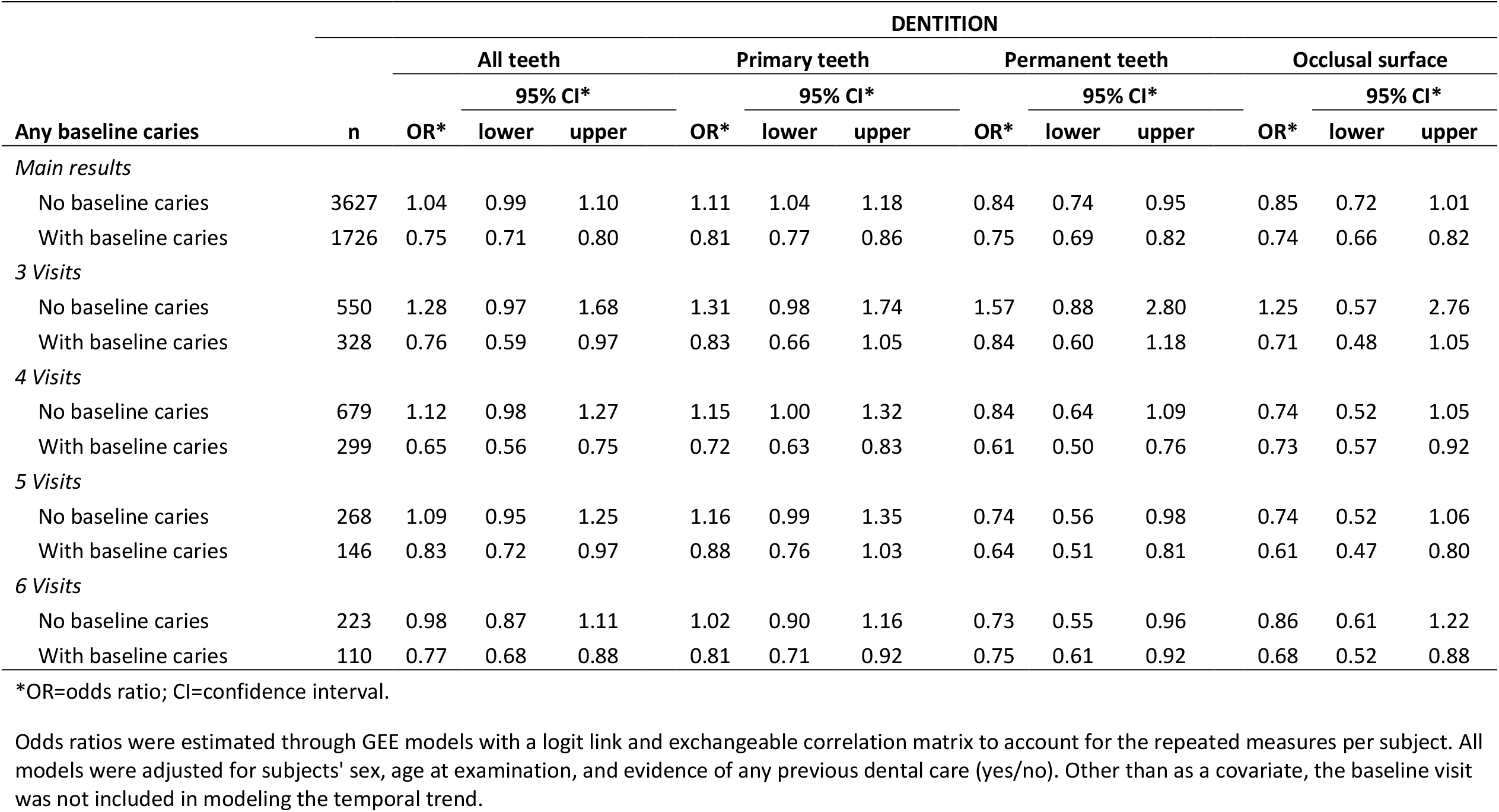
Temporal trend in the presence of untreated caries over visits: Restriction to fixed numbers of visits to explore attrition bias in all 33 schools (Phases I and II).

**eTable 4b.**
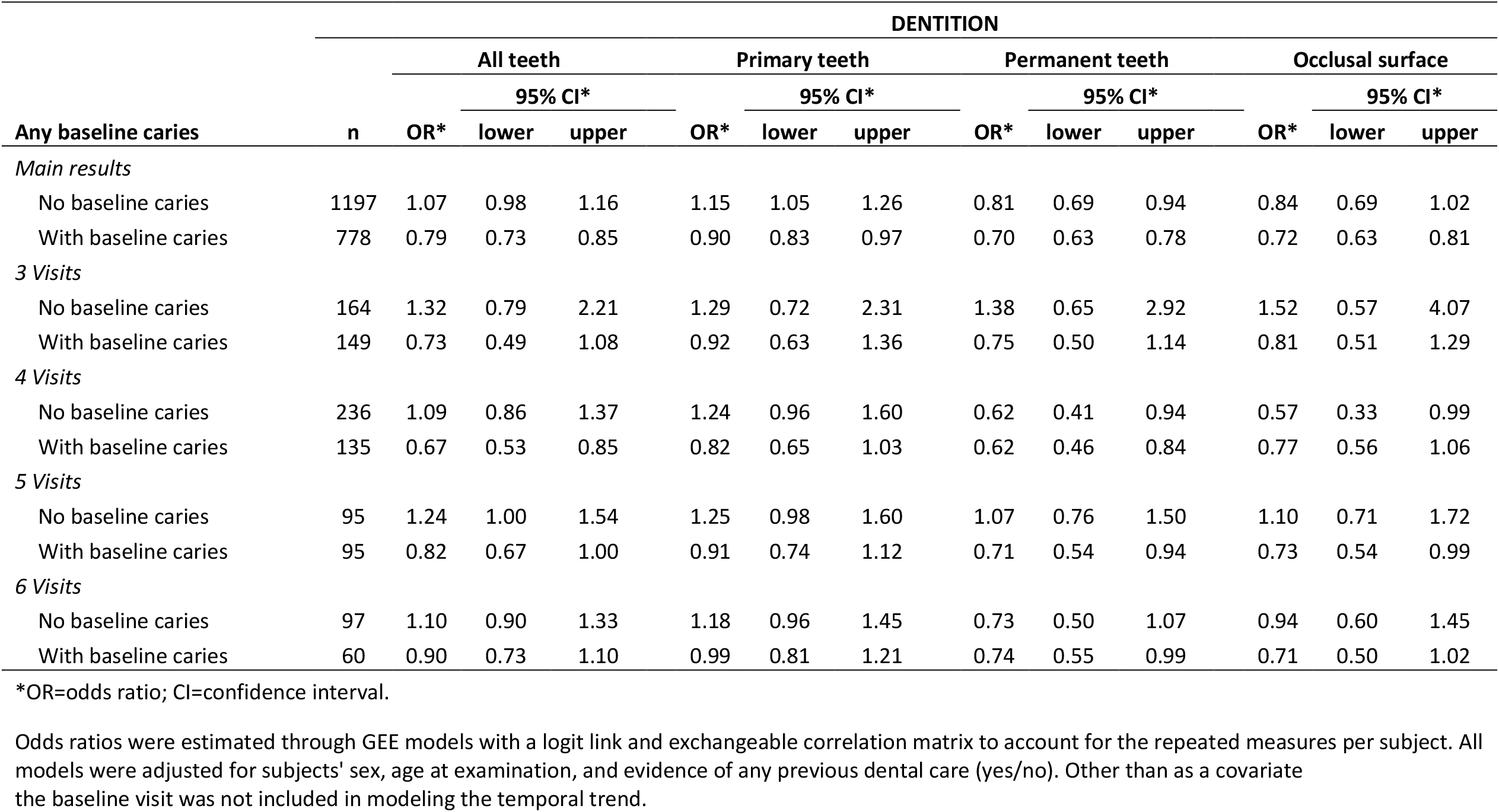
Temporal trend in the presence of untreated caries over visits: Restriction to fixed numbers of visits to explore attrition bias in first 6 schools (Phase I).

**eTable 4c.**
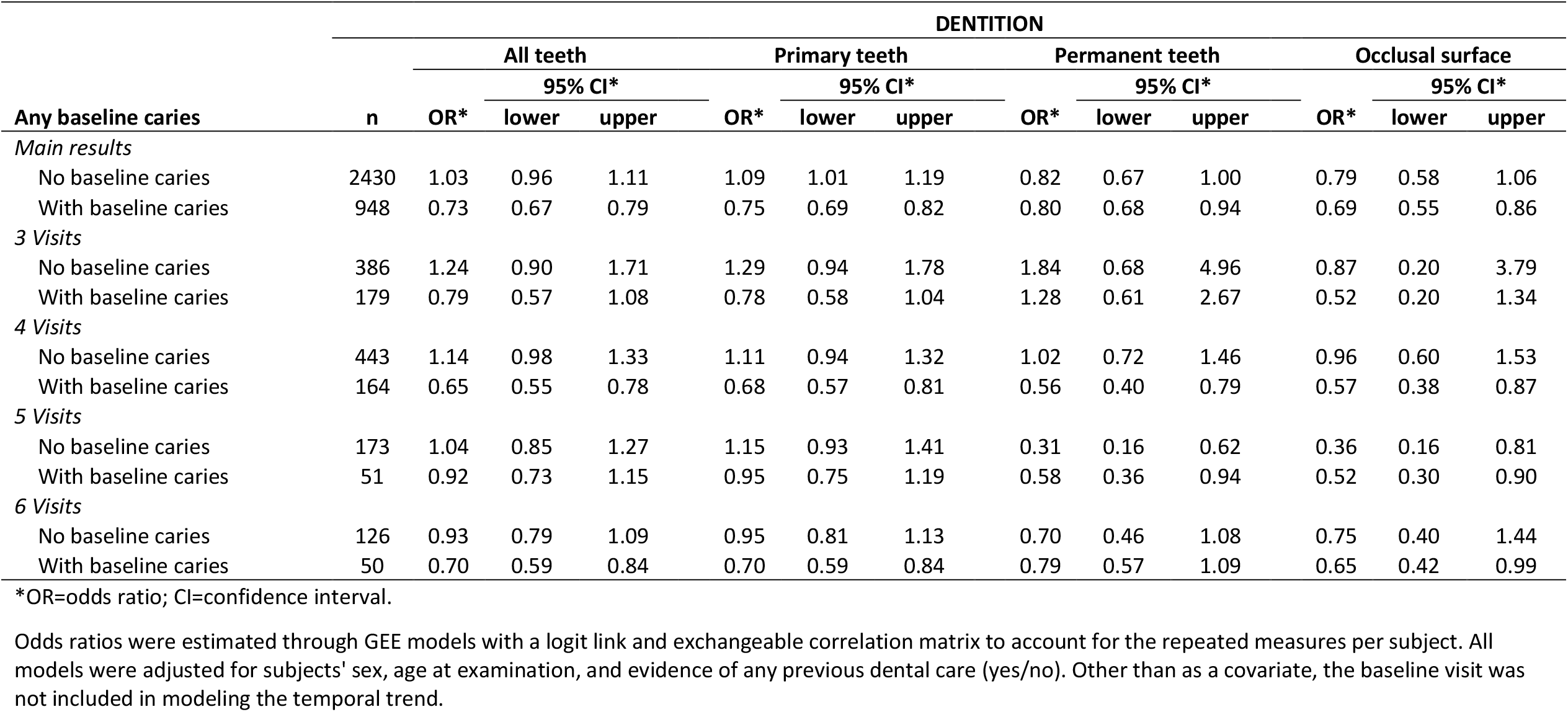
Temporal trend in the presence of untreated caries over visits: Restriction to fixed numbers of visits to explore attrition bias in later 27 schools (Phase II).

**Supplemental information 5: Sensitivity analyses to address other possible sources of bias**.

The Investigational Review Board approval and approval from the schools allowed for the collection of only minimal demographic and health information from study subjects, which limited the extent to which we could directly account for possible confounders and sources of bias. We considered that either the treatment effect or odds of caries might be different in children who entered the study with more caries (i.e. more affected sites) at baseline. The study subjects who entered at the youngest ages had the highest possibility for longer follow-up. Moreover, the numbers of subjects decreased over time.

To address these various concerns, we repeated the primary analyses after restricting to subjects who a) had <4 teeth with any untreated caries at baseline, b) had <6 teeth with any treated or untreated caries at baseline, or c) were younger than 8 years at baseline. We also repeated analyses, excluding visits higher than five.

Estimated odds ratios changed only slightly throughout these analyses in all schools combined (Table 5a), in Phase I (Table 5b), and Phase II (Table 5c).

**eTable 5a.**
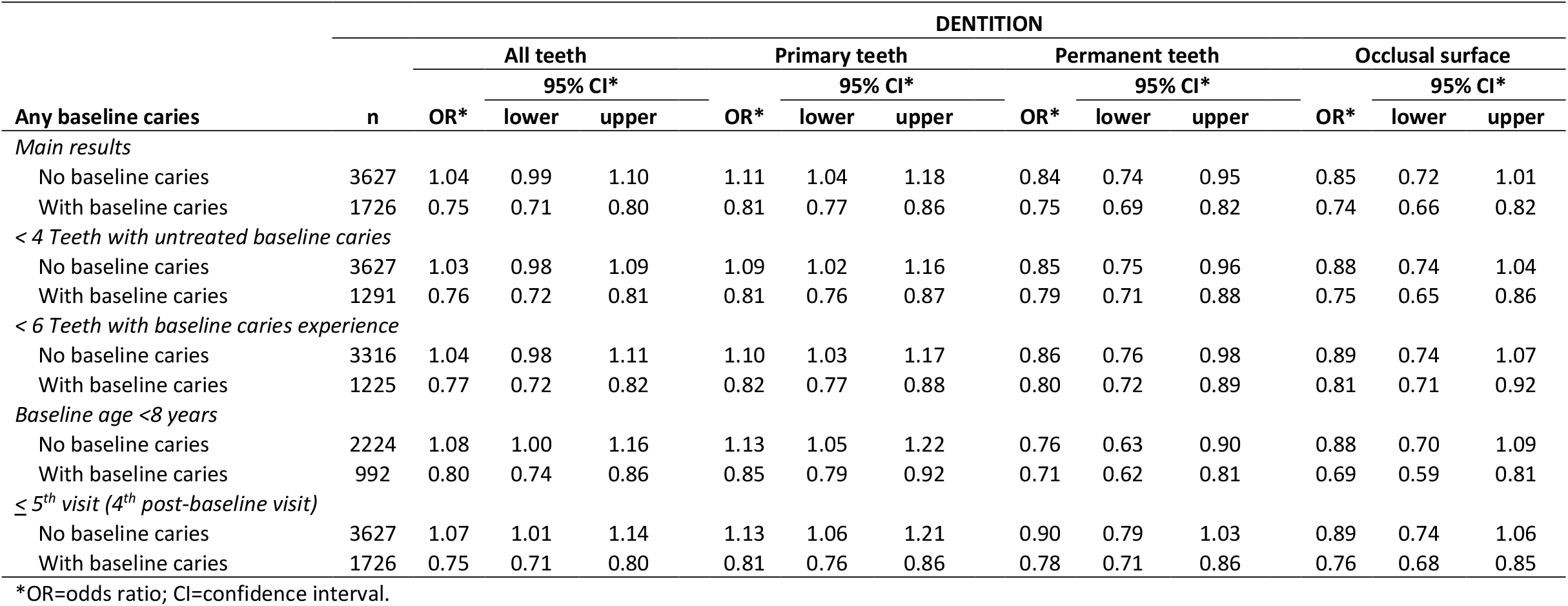
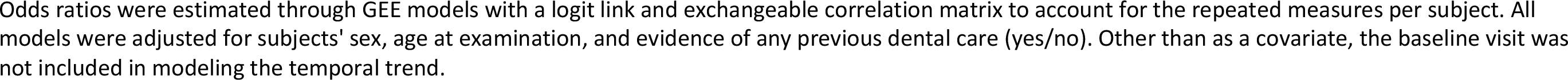
Temporal trend in the presence of untreated caries over visits: Exploration of other sources of bias in all 33 schools (Phases I and II).

**eTable 5b.**
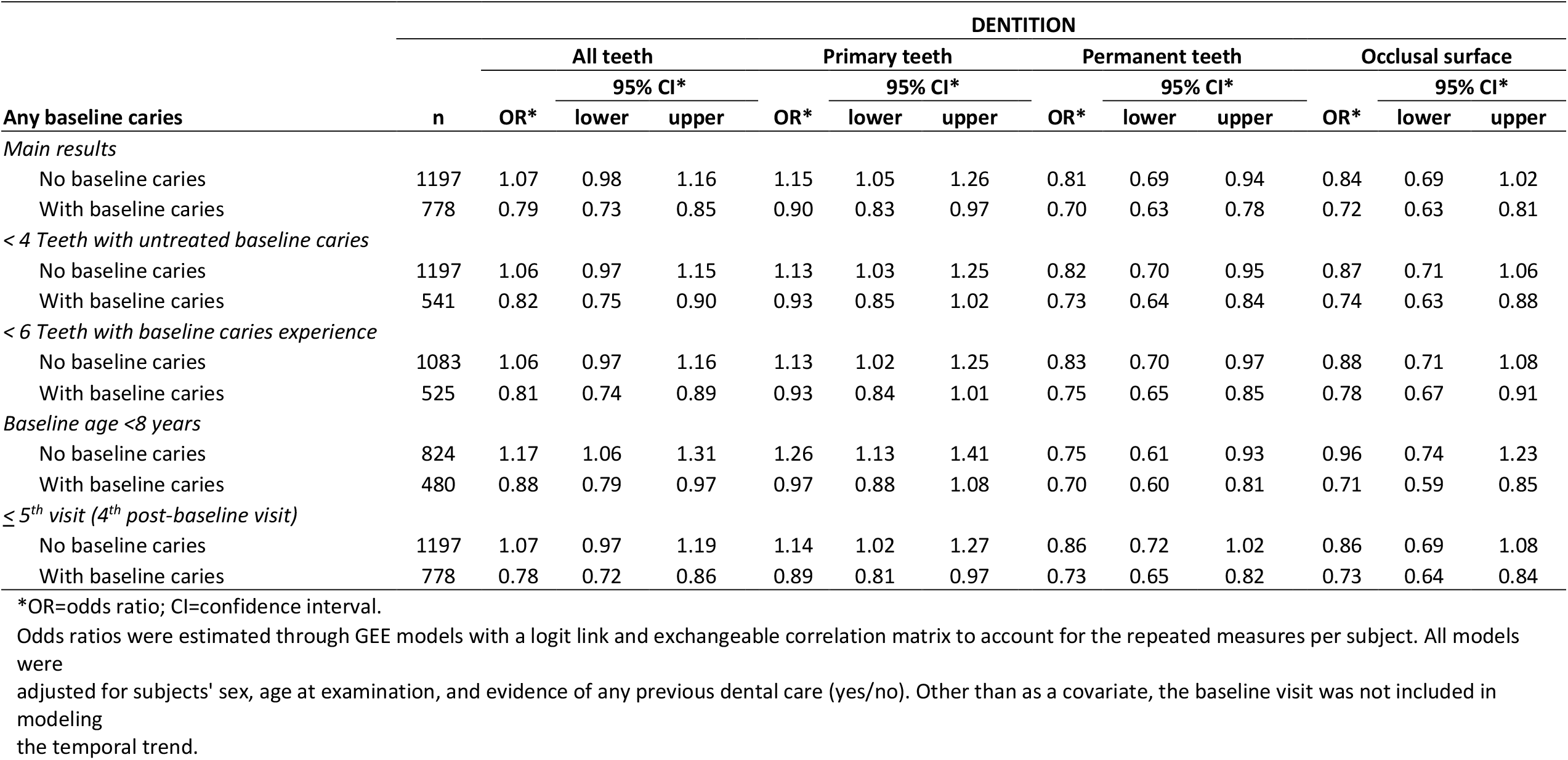
Temporal trend in the presence of untreated caries over visits: Exploration of other sources of bias in the first 6 schools (Phase I).

**eTable 5c.**
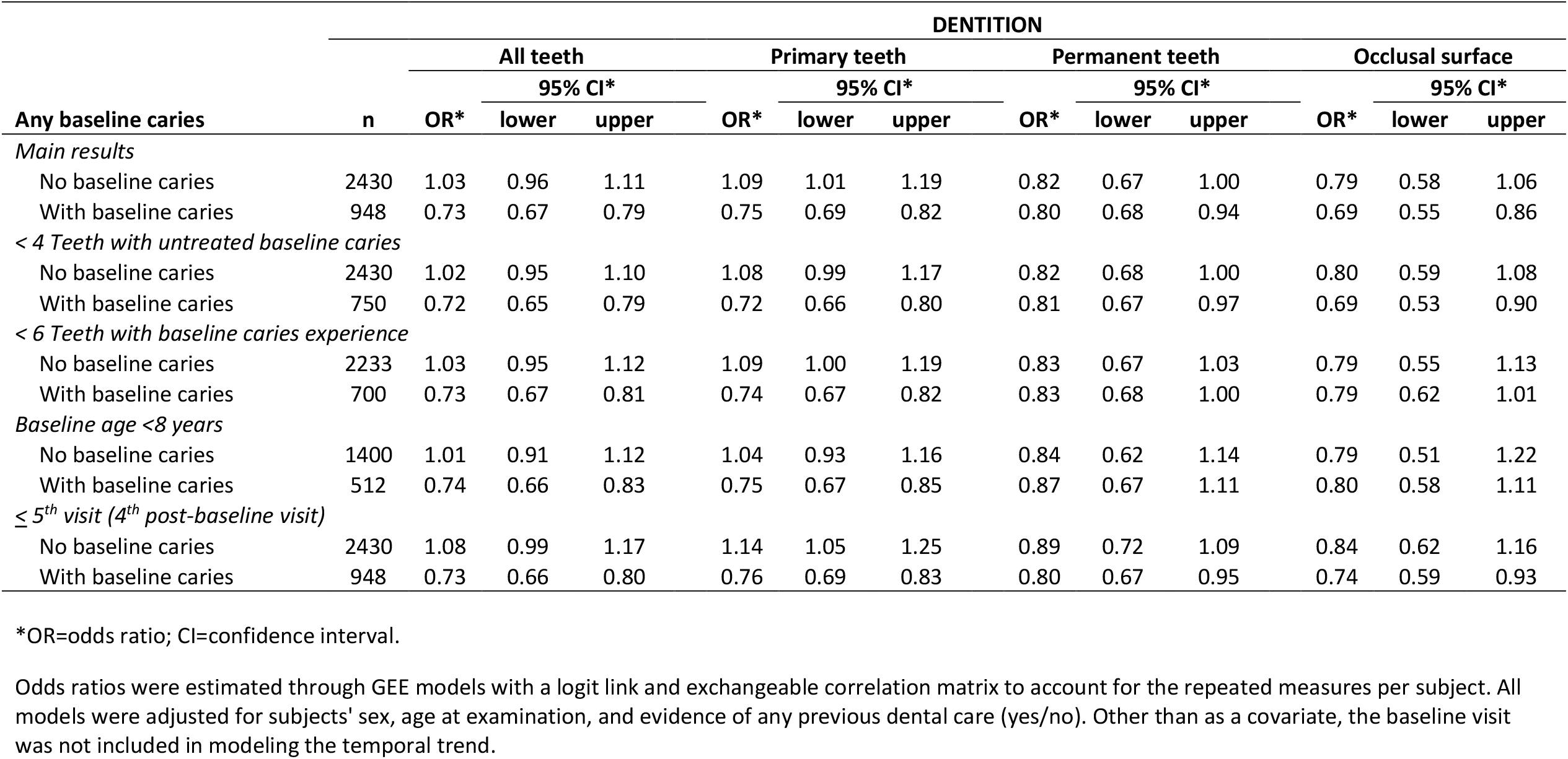
Temporal trend in the presence of untreated caries over visits: Exploration of other sources of bias in later 27 schools (Phase II).

**Supplemental information 6:** Strengthening the Reporting of Observational Studies in Epidemiology

|  | Item No | Recommendation | Page No |
| --- | --- | --- | --- |
| Title and abstract | 1 | (a) Indicate the study’s design with a commonly used term in the title or the abstract | 1 |
|  |  | (b) Provide in the abstract an informative and balanced summary of what was done and what was found | 3 |
| Introduction |  |  |  |
| Background/rationale | 2 | Explain the scientific background and rationale for the investigation being reported | 4 |
| Objectives | 3 | State specific objectives, including any prespecified hypotheses | 4 |
| Methods |  |  |  |
| Study design | 4 | Present key elements of study design early in the paper | 6 |
| Setting | 5 | Describe the setting, locations, and relevant dates, including periods of recruitment, exposure, follow-up, and data collection | 6 |
| Participants | 6 | (a) Give the eligibility criteria, and the sources and methods of selection of participants. Describe methods of follow-up | 6 |
|  |  | (b) For matched studies, give matching criteria and number of exposed and unexposed | 6 |
| Variables | 7 | Clearly define all outcomes, exposures, predictors, potential confounders, and effect modifiers. Give diagnostic criteria, if applicable | 7 |
| Data sources/measurement | 8* | For each variable of interest, give sources of data and details of methods of assessment (measurement). Describe comparability of assessment methods if there is more than one group | 7, 8 |
| Bias | 9 | Describe any efforts to address potential sources of bias | 8 |
| Study size | 10 | Explain how the study size was arrived at | 7 |
| Quantitative variables | 11 | Explain how quantitative variables were handled in the analyses. If applicable, describe which groupings were chosen and why | 7 |
| Statistical methods | 12 | (a) Describe all statistical methods, including those used to control for confounding | 7 |
|  |  | (b) Describe any methods used to examine subgroups and interactions | 8 |
|  |  | (c) Explain how missing data were addressed | 8 |
|  |  | (d) If applicable, explain how loss to follow-up was addressed |  |
|  |  | (e) Describe any sensitivity analyses | 8, 9 |
| Results |  |  |  |
| Participants | 13* | (a) Report numbers of individuals at each stage of study—eg numbers potentially eligible, examined for eligibility, confirmed eligible, included in the study, completing follow-up, and analysed | 10 |
|  |  | (b) Give reasons for non-participation at each stage | 12 |
|  |  | (c) Consider use of a flow diagram | NA |
| Descriptive data | 14* | (a) Give characteristics of study participants (eg demographic, clinical, social) and information on exposures and potential confounders | 10 |
|  |  | (b) Indicate number of participants with missing data for each variable of interest | 11 |
|  |  | (c) Summarise follow-up time (eg, average and total amount) | 11, 12 |
| Outcome data | 15* | Report numbers of outcome events or summary measures over time | 10 |
| Main results | 16 | (a) Give unadjusted estimates and, if applicable, confounder-adjusted estimates and their precision (eg, 95% confidence interval). Make clear which confounders were adjusted for and why they were included<br>(b) Report category boundaries when continuous variables were categorized<br>(c) If relevant, consider translating estimates of relative risk into absolute risk for a meaningful time period | 18,<br>21,<br>29-41<br>21 |
| Other analyses | 17 | Report other analyses done—eg analyses of subgroups and interactions, and sensitivity analyses | 28-41 |
| **Discussion** |  |  |  |
| Key results | 18 | Summarise key results with reference to study objectives | 12 |
| Limitations | 19 | Discuss limitations of the study, taking into account sources of potential bias or imprecision. Discuss both direction and magnitude of any potential bias | 12-13 |
| Interpretation | 20 | Give a cautious overall interpretation of results considering objectives, limitations, multiplicity of analyses, results from similar studies, and other relevant evidence | 13-14 |
| Generalisability | 21 | Discuss the generalisability (external validity) of the study results | 15 |
| **Other information** |  |  |  |
| Funding | 22 | Give the source of funding and the role of the funders for the present study and, if applicable, for the original study on which the present article is based | 17 |

